# Network alterations in temporal lobe epilepsy during non-rapid eye movement sleep and wakefulness

**DOI:** 10.1101/2023.10.06.23296655

**Authors:** I Rigoni, BJ Vorderwülbecke, M Carboni, N Roehri, L Spinelli, G Tononi, M Seeck, L Perogamvros, S Vulliémoz

**Affiliations:** EEG and Epilepsy unit, University Hospital and Faculty of Medicine of Geneva, University of Geneva, Switzerland; Epilepsy-Center Berlin-Brandenburg, Department of Neurology with Experimental Neurology, Charité - Universitätsmedizin Berlin, corporate member of Freie Universität Berlin and Humboldt-Universität zu Berlin, Berlin, Germany; Department of Psychiatry, University of Wisconsin, Madison, WI, USA; Center for Sleep Medicine, Department of Psychiatry, University Hospitals of Geneva, Geneva, Switzerland

**Keywords:** Temporal lobe epilepsy, functional connectivity, EEG, NREM sleep stage 2, brain networks

## Abstract

**Objective:** Investigate sleep and temporal lobe epilepsy (TLE) effects on EEG-derived brain networks.

**Methods:** High-density EEG was recorded during non-REM sleep (N2) and wakefulness in 23 patients and healthy controls (HC). Epochs without epileptic discharges were source-reconstructed in 72 brain regions and connectivity was estimated. Network integration (Efficiency, E) and segregation (Clustering Coefficient, CC) at global and hemispheric level (GE, avgCC, HE, HCC) were calculated. These were compared between groups across frequency bands and correlated with the individual proportion of wakefulness-or sleep-related seizures.

**Results:** Patients had higher delta GE, delta avgCC and theta avgCC than controls, irrespective of the vigilance state (TLE > HC, p<.01). During wakefulness, theta GE of patients was higher than controls (p<.001) and, for patients, theta GE during wakefulness was higher than during N2 (p<.05). Wake-to-sleep differences in TLE were notable only in the ipsilateral hemisphere (HE and HCC, p<.05). Only measures from wakefulness recordings correlated with the proportion of wakefulness-or sleep-related seizures.

**Conclusions:** TLE network alterations are more prominent during wakefulness and at lower frequencies. Increased integration *and* segregation suggest a pathological ‘small world’ configuration with a possible inhibitory role.

**Significance:** Network alterations in TLE occur and are easier to detect during wakefulness.

## 1 Introduction

Epilepsy is the brain’s enduring predisposition to generate epileptic seizures due to paroxysmal, abnormal excessive or synchronous neuronal activity (Fisher et al., 2014). In 70% of patients, epilepsy is focal (Bosak et al., 2019). This means that seizures originate in networks limited to one hemisphere of the brain, although these networks can be widely distributed (Berg et al., 2010; Fisher et al., 2017). Depending on the specific networks involved, clinical presentation of seizures varies individually, and so does their interplay with the sleep-wake cycle. Depending on seizure type and epilepsy syndrome, seizures may occur predominantly or exclusively either during wakefulness or during sleep (Bazil, 2019; Jain and Kothare, 2015). While rapid eye movement (REM) sleep stage appears protective against seizures, most sleep-related seizures occur during non-REM sleep stages 1 and 2 (N1 and N2) (Bazil, 2019; Jain and Kothare, 2015). IEDs occur more frequently during sleep than during wakefulness, and more frequently during non-REM sleep than during REM sleep (Malow et al., 1999, 1998). On the other hand, IEDs during REM sleep appear more spatially restricted and more helpful to localize the epileptic focus than during non-REM sleep (Sammaritano et al., 1991; Yuan and Sun, 2020). In sum, sleep is a modulator of epilepsy, and their interaction is multifaceted.

Epileptic networks have been studied mainly during wakefulness, and results suggest that connectivity analyses could help differentiate patients with temporal lobe epilepsy (TLE) from healthy controls (HC) (Verhoeven et al., 2018), localize the seizure onset zone (Staljanssens et al., 2017) and predict seizure freedom after surgery (Varatharajah et al., 2022). Recent results suggest that high integration -meaning a more efficient information transfer across brain regions-could be a biomarker of epilepsy: during segments of scalp electroencephalography (EEG) without IEDs, TLE patients showed more integrated brain networks than HC (Carboni et al., 2020). Moreover, patients with poorer surgery outcome showed higher functional connectivity between the propagation zone and the non-involved zone (Lagarde et al., 2018) and higher integration, as measured by the global efficiency (GE) of the IED network (Carboni et al., 2019). Despite these encouraging findings, functional connectivity analyses have not made their way into clinical practice yet (van Mierlo et al., 2014), likely due to the non-convergence of results from different studies (Slinger et al., 2021).

Surprisingly, connectivity studies on epilepsy during sleep are much rarer. When looking at IED during wakefulness and NREM sleep, it was found that IED co-occurrence between brain regions increases during NREM sleep, especially in neocortical regions (Lambert et al., 2018). Given that medial temporal lobe structures have a higher propensity to generate spike even if they are not part of the seizure onset zone (Lambert et al., 2018), these results suggest that the IED-propagation network from medial temporal structures to the neocortex is wider than in wakefulness. When using cortico-cortical evoked potentials as proxies of effective connectivity, it was shown that the connectivity between both epileptogenic and non-epileptogenic structures as well as brain excitability in general increased during NREM sleep (Arbune et al., 2020; Usami et al., 2015). Besides these few findings, the effects of *wakefulness vs. sleep* on brain network characteristics in *patients with epilepsy vs. healthy controls* have not yet been studied systematically, and we aimed at filling this gap.

Based on previous findings (Carboni et al., 2020, 2019), we hypothesized that (1) network integration is higher in TLE patients than in HC subjects; (2) network alterations between wakefulness and non-REM sleep are most pronounced in structures affected by the epilepsy as compared to the contralateral side; (3) the extent of network alterations between wakefulness and N2 sleep is coupled to the individual predisposition towards wakefulness-or sleep-related seizures. In addition to efficiency as a measure of network integration, we aimed at studying the clustering coefficient (CC) as a measure of network segregation.

## 2 Methods

### 2.1 Participants

TLE patients were retrospectively selected from the database of the EEG and Epilepsy Unit at University Hospitals Geneva, based on the following criteria: age >18 years, diagnosis of TLE, available 257-channel EEG recording of at least 60 minutes, intact skull and no extratemporal brain lesion on the magnetic resonance imaging (MRI) scan. HC were retrospectively recruited at the University of Wisconsin–Madison, for another sleep research project (Perogamvros et al., 2017; Siclari et al., 2017), according to the following criteria: older than 25 years old, with no insomnia symptoms and BMI <35 kg/m2. TLE patients and HC were matched according to age and sex. In TLE patients, based on all available information from patient history and video-EEG findings, proportions of wake-vs. sleep-related seizures were graded in 20%-steps.

Among 511 patients with epilepsy screened, 41 met the inclusion criteria. Of these, 14 were excluded because their EEG recordings did not contain enough N2 sleep, and another 4 were excluded because they could not be age-matched with our control database. Eventually, 23 TLE patients (median age 36 years, range 24-60, 12 females, 8 left TLE, 13 right TLE, 2 bilateral TLE) and 23 HC (median age 36 years, range 24-61, 12 females) were retained for analyses. For individual clinical details, see Table 1. Resting state wakefulness and N2 sleep EEG epochs were available for all TLE patients and HC, except that 10 HC had no wake EEG. The study was carried out in accordance with the declaration of Helsinki. For TLE patients, written informed consent was waived for the reuse of clinical routine data. All healthy control subjects gave written informed consent. Ethical approval was granted by the ethics board of the District of Geneva and the institutional review board at the University of Wisconsin–Madison.

**Table 1:** Patient clinical details. Abbreviations: M, male; F, female; L, left; R, right; B, bi-temporal; HS, hippocampal sclerosis; FCD, focal cortical dysplasia; OXC, Oxcarbazepine; CBZ, Carbamazepine; VPA, Valproic acid; LEV, Levetiracetam; TPM, Topiramate; LTG, Lamotrigine; ZNS, Zonisamide; CLB, Clobazam; PGB, Pregabalin; LCM, Lacosamide; PER, Perampanel

| ID_TLE | Sex | Age when epilepsy started (range) | Duration of disease at recording (years) | Percentage of seizures during wakefulness | TLE side | Etiology | ILAE at 12 months | ASM at evaluation |
| --- | --- | --- | --- | --- | --- | --- | --- | --- |
| P1 | M | 21-25 | 4 | 0-20% | R | HS | 1 | OXC |
| P2 | M | 21-25 | 6 | 100% | L | HS + atrophy of the L mamillary body and L temporal lobe | 1 | CBZ, VPA |
| P3 | M | 6-10 | 20 | 40-60% | L | (unknown) | 3 | LEV, TPM |
| P4 | M | 11-15 | 14 | 40-60% | R | Cavernoma | 1 | OXC, VPA |
| P5 | M | 21-25 | 6 | 100% | R | HS + asymmetry of mamillary body | 1 | CBZ |
| P6 | M | 16-20 | 17 | 0-20% | R | (unknown) | 4 | LTG, ZNS |
| P7 | M | 0-5 | 32 | 40-60% | R | HS | 1 (6mo) | CLB, LTG, VPA |
| P8 | M | 31-35 | 6 | 40-60% | R | FCD or glioma, unclear (MR only) | NA | OXC |
| P9 | M | 16-20 | 27 | 100% | L | HS (MR only) | 1 | CBZ, LTG, TPM |
| P10 | M | 36-40 | 10 | 60-80% | B | HS (MR only) | NA | LEV, LTG, VPA |
| P11 | M | 26-30 | 24 | 100% | L | HS | 1 | CBZ, CLB, LCM, LEV, LTG |
| P12 | F | 6-10 | 14 | 40-60% | L | amygdaloidal origin, unclear | NA | VPA |
| P13 | F | 21-25 | 7 | 40-60% | R | (unknown) | 5 | OXC, PGB |
| P14 | F | 16-20 | 14 | 80-100% | L | (unknown) | NA | LEV, LCM |
| P15 | F | 21-25 | 5 | 100% | R | Dysplasia | 1 | ZNS |
| P16 | F | 26-30 | 6 | 100% | L | (unknown) | NA | LTG, PGB |
| P17 | F | 26-30 | 8 | 100% | L | ganglioglioma I° | 1 | ZNS |
| P18 | F | 11-15 | 25 | 40-60% | B | HS | NA | LTG, ZNS |
| P19 | F | 36-40 | 7 | 100% | R | hippocampal gliosis | 1 | LTG, VPA, ZNS |
| P20 | F | 26-30 | 18 | 60-80% | R | hippocampal gliosis | 1 (6mo) | LEV, LTG, ZNS |
| P21 | F | 21-25 | 26 | 20-40% | R | (unknown) | NA | PER, TPM, VPA |
| P22 | F | 21-25 | 27 | 60-80% | R | (unknown) | 3 | CBZ, TPM |
| P23 | F | 21-25 | 35 | 40-60% | R | HS | 1 | LEV |

### 2.2 Data acquisition and preprocessing

High-density EEG recordings of patients (257 electrodes, Electrical Geodesics Inc., now Magstim EGI, Eugene, OR; sampling rate 250-1000 Hz) were acquired in the context of pre-surgical evaluation, sometimes overnight. A board-certified neurologist (BJV) extracted IED-free segments of wakefulness and of stable, *i*.*e*. arousal-free, non-rapid eye movement (NREM) sleep stage 2 (N2) according to the standard criteria (Iber et al., 2007). For wake EEG, task-free wakefulness was chosen, most often with closed eyes.

High-density EEG was recorded from HC (257 electrodes, Electrical Geodesic Inc. sampling rate = 500 Hz) with a paradigm including serial awakenings from across the night, as reported in (Perogamvros et al., 2017; Siclari et al., 2017). Supplementary electrodes were used to monitor eye movements and to record submental electromyogram. Sleep scoring was performed over 30-second epochs according to standard criteria (Iber et al., 2007), and segments of N2 sleep were extracted. For wake EEG, participants were fixating a cross as described in (Perogamvros et al., 2017) and segments without artefacts (i.e. eye -blinks) were chosen for the analyses. A bandpass Butterworth filter was used to filter the data between 1 and 35 Hz and a notch Butterworth filter was applied at 50 Hz. Data were down-sampled to 250 Hz; sensors with a poor signal-to-noise ratio were visually identified and removed. Sensors placed on the cheeks and neck were removed, leaving a maximum of 204 channels for the forward and inverse solution (Vorderwülbecke et al., 2020). Average re-referencing was applied, and 20 epochs of 2 seconds were extracted for each participant and state (wake vs. N2), where available (the required minimum was 15 epochs). Data containing K complexes was not considered for epoch selection to reduce variability between the selected epochs.

### 2.3 Source reconstruction

The MNI Colin-27 head served as a template for the source space reconstruction (Holmes et al., 1998). Based on 1 mm3 isotropic resolution, a 3D cerebral grey matter mask was generated with Freesurfer (version 6.0.1) (Reuter et al., 2012). The grey matter mask was parcelled into 82 regions of interest (ROI) based on Lausanne 1 scale parcellation (Cammoun et al., 2012; Hagmann et al., 2008), using the open-source Connectome Mapper 3 (Tourbier et al., 2020). Using Cartool 3.80 (version 6164) (Brunet et al., 2011), a regular 3D grid containing 5,020 solution points was distributed throughout the grey matter mask. The Locally Spherical Model with Anatomical Constraints (LSMAC) served as forward model, and the Local Autoregressive Average (LAURA) as inverse model (Michel and Brunet, 2019). By excluding the subcortical grey matter (thalamus, caudate, putamen, pallidum and accumbens area), the source space was restricted to 72 cortical ROIs. The time-series of the 72 ROI were reconstructed as the first singular vector computed by a singular-value decomposition of all the 3D source-time courses in the same ROI (Rubega et al., 2019).

### 2.4 Connectivity

We quantified the statistical dependencies between the 72 brain areas with the debiased weighted phase lag index (dwPLI), using the Matlab-based Fieldtrip toolbox (Oostenveld et al., 2011). In previous studies, we used metrics based on Granger-causal modelling (Carboni et al., 2020; Coito et al., 2016). However, since we compare vigilance states (N2 sleep and wakefulness) known to exhibit distinct frequency content, we chose an unbiased metric that could account for signal variations across different frequency bands. The resulting connectivity matrix (72 x 72 x 81, where 81 is the number of frequency bin spacing from 0 to 40 Hz) was averaged across the different frequency bands. This yielded five average matrices for each participant in each vigilance state: delta (1-4 Hz), theta (4-8 Hz), alpha (8-12 Hz), sigma (12-16 Hz) and beta (16-30 Hz).

### 2.5 Graph analysis

The connectivity matrices describe the brain as a network where the nodes are the different ROIs and the edges are their statistical dependencies. To characterize the *integration* and *segregation* of brain networks, we extracted the global efficiency (GE) and the average clustering coefficient (avgCC), respectively (Rubinov and Sporns, 2010). Briefly, the GE reflects how well information is propagated over distributed brain regions and the avgCC reflects the preponderance of clustered connectivity around individual nodes. They are the average of the nodal measures (*E*_*i*_ and *CC*_*i*_) and are formally calculated as follows:

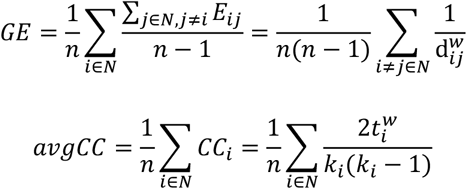

where 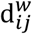 is the distance between node *i* and node *j* measured as the sum of the inverse of the strongest weights that directly connect *i* and *j, k*_*i*_ is the degree of the node and 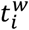 is the number of triangles attached to the node (Latora and Marchiori, 2001; Rubinov and Sporns, 2010). These metrics were also calculated at the hemispheric level and at the level of the temporal lobes, yielding hemispheric efficiency and CC (HE and HCC) and temporal efficiency and CC (TE and TCC). More details are provided in the Appendix. For statistical analyses, we investigated the hemisphere ipsilateral to the epileptic focus and the contralateral one.

### 2.6 Statistics

We modelled the global metrics, GE and avgCC, for each of the 5 frequency bands individually with a linear mixed model (LMM). The LMM was chosen to account for missing values -HC for whom the recordings during wakefulness were not available (N_HC,N2_=23, N_HC,W_=13, N_TLE,N2_=23, N_TLE,W_=23). Group (PAT vs HC) and vigilance state (wakefulness vs N2 sleep) were modelled as fixed effects while subjects were modelled as random effects [*Y* ∼ *group* ∗ *state* + (1|*ID*)]. GE and avgCC were log-transformed and the model was fitted. Then, we ran a Type-II ANalyses Of VAriance (ANOVA) of the mixed effects using F-tests and the significance of the interaction (or main effect of Group and State) was Bonferroni-adjusted for the 5 tests ran (one for each frequency band). When significant interactions were found, a further Bonferroni correction was applied to account for the 4 post-hoc tests. Multilevel analyses were conducted in RStudio, version 2022.07.1, with R statistical language, version 4.1.2 (RStudio Team, Boston, MA, 2019).

Then we looked at how graph measures changed, between vigilance states, in the ipsilateral and contralateral hemisphere (and temporal lobe). Results on HC are presented in the Appendix. For each hemisphere and frequency band, we compared the hemispheric and temporal graph metric (HE and HCC, and TE and TCC respectively) in N2 vs. wake with a two-sided Wilcoxon signed-rank test. A false-discovery rate (FDR) correction was applied to correct for the 5 tests. For hemispheric and temporal-lobe analyses, we retained only the HC for whom we had recordings during both wake and sleep (N=13) and the patients who had either right or left TLE (N=21), but not bilateral (N=2). To avoid an effect of intrinsic physiological difference between the right and left hemisphere, the right and left hemispheres were flipped in a proportion of the HC (5/13=38%) corresponding to the proportion of LTLE (8/21=38%).

In the TLE population, correlation analyses were run between the proportion of wake-related seizures and the network metrics (efficiency and clustering coefficient) calculated at the global, hemispheric and temporal level. Correlations were only performed for the frequency bands where significant differences emerged. A Spearman correlation test was chosen for its smaller sensitivity to outliers. Both FDR-corrected and uncorrected p values are reported.

Finally, when the analyses on the global network metrics (GE or avgCC) resulted in a significant interaction, we ran an *edge-wise analysis* on the connectivity matrices. We used the network-based statistic (NBS) method to identify any subnetwork that significantly differed between the two groups compared: more details are provided in the Appendix.

Lastly, correlation between the power of the source-reconstructed signal and the global network metrics is provided in the Appendix.

## 3 Results

### 3.1 Global network metrics

#### 3.1.1 Global efficiency

In delta, a significant main effect of group [F_(1,45.524)_= 15.38, p= 0.0015] shows that GE was higher in TLE than in controls (Fig. 1, first row). In sigma, a significant main effect of vigilance [F_(1, 38.069)_= 16.93, p= 0.001] shows that GE during N2 sleep is larger than during wakefulness. In theta, a significant group x vigilance interaction [F_(1, 42.611)_= 8.08, p= 0.0342] is found and post-hoc tests show that GE was higher in TLE than in HC during wakefulness [t_(77.5)_= -4.22, p=0.0003] and that, in TLE, GE is higher during wakefulness than N2 sleep [t_(36.6)_= -2.85, p=0.0285].

**Fig 1:**
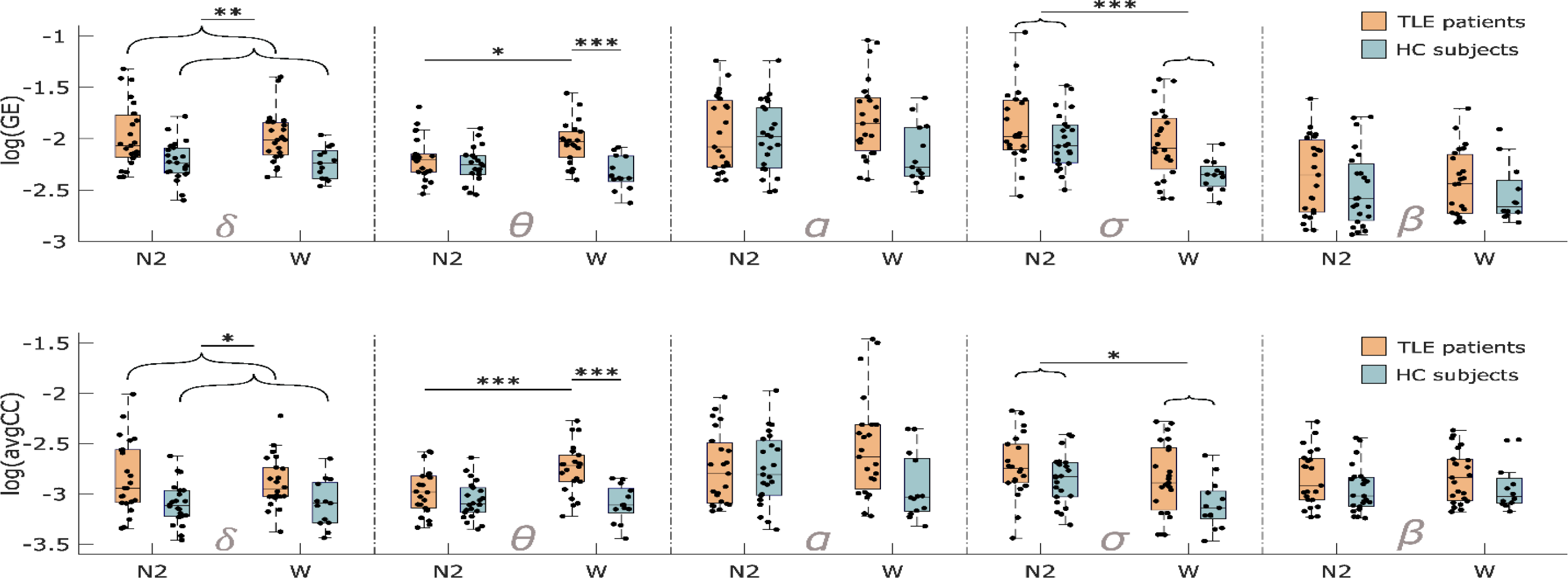
Results of the LMM analyses performed, individually, for each frequency band on: a) GE and b) avgCC. For each graph measure, the five frequency bands are reported. The figure reports the main effect of group in delta band (first column) and in theta band for avgCC (second column); the significant group x vigilance interaction in theta band for GE (second column); the main effect of vigilance in sigma band (fourth column). The asterisks indicate statistical significance (* for p<.05, ** for p<.01 and *** for p<.001).

#### 3.1.2 Average clustering coefficient

In delta, a significant main effect of group [F_(1,45.246)_= 11.49, p= 0.0073] shows that avgCC was higher in TLE than in controls (Fig. 1, second row). In sigma, a significant main effect of vigilance [F_(1,38.49)_= 13.74, p= 0.0033] shows that avgCC is higher during N2 sleep than during wakefulness. In theta, the group x vigilance interaction did not reach significance [F_(1,43.753)_= 6.7, p= 0.0651], but a main effect of the group [F_(1,44.707)_= 11.97, p= 0.006] shows that avgCC was higher in TLE than in controls.

#### 3.1.3 Correlation with seizure preponderance

Correlation analyses were run for delta, theta and sigma bands (NTLE=23). They show that the proportion of wake-related seizures correlates with global network metrics only during wakefulness and only in delta. A positive linear correlation was found for delta GE measured during wakefulness (delta GE: rho=0.41, puncorr 0.05, pFDRcorr= 0.17), meaning that for patients with high delta GE during wakefulness most of the seizures occur during the wakefulness. Instead, no correlation was observed during N2 sleep. Similarly, delta avgCC measured during wakefulness positively correlated with the percentage of wakefulness-related seizures (delta avgCC: rho=0.45, puncorr 0.03, pFDRcorr= 0.2).

### 3.2 Hemispheric network metrics

We then investigated whether the wake-to-sleep changes in network efficiency of TLE were lateralized toward the hemisphere ipsilateral or contralateral to the epileptic focus.

#### 3.2.1 Hemispheric efficiency

We found significant changes of HE between wakefulness and sleep only in the ipsilateral hemisphere: HE dropped from wakefulness to N2 sleep in theta band (z=2.6, p=.0217) and increased in sigma (z=-2.8, p=.0217, see Fig. 2a). No change was observed in the other bands nor in any band in the contralateral hemisphere, nor in HC (Fig. S1a).

**Fig 2:**
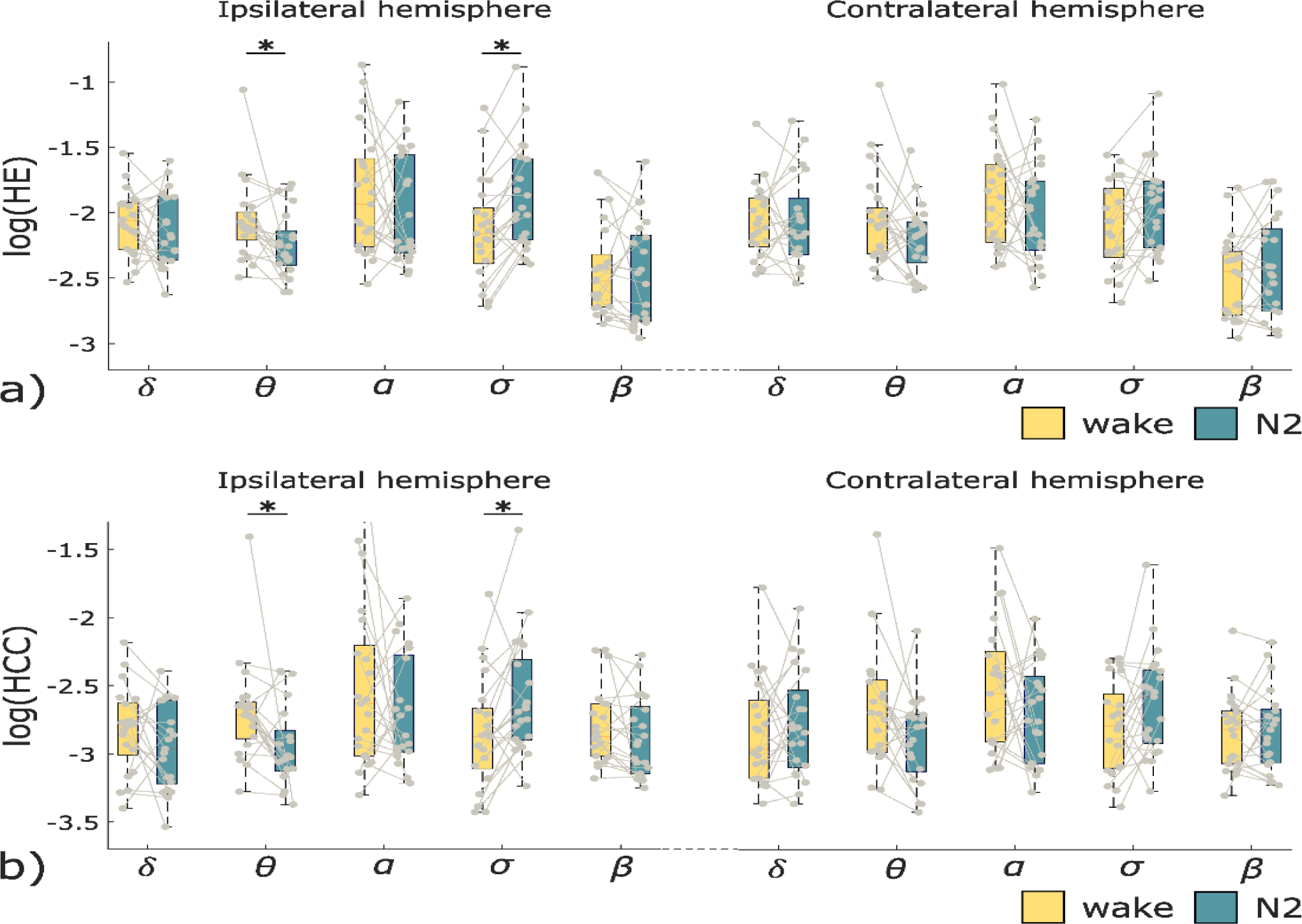
Results of the Wilcoxon tests run on the hemispheric network metrics of TLE patients during wakefulness and N2 sleep. a) Hemispheric efficiency (HE) during wakefulness (yellow) and N2 sleep (green) in the hemisphere ipsilateral and contralateral to the epileptic focus. b) Hemispheric clustering coefficient (HCC) during wakefulness (yellow) and N2 sleep (green) in the hemisphere ipsilateral and contralateral to the epileptic focus. The asterisks indicate statistical significance (p<.05).

#### 3.2.2 Hemispheric clustering coefficient

Similarly, we found significant changes of HCC between wakefulness and sleep only in the ipsilateral hemisphere: HCC decreased from wakefulness to N2 sleep in the theta band (z=2.7, p=.0353) and increased in sigma (z=-2.3, p=.0474, see Fig 2b). No change was observed in the other bands nor in any band in the contralateral hemisphere, nor in healthy controls (Fig. S1b).

#### 3.3.3 Correlation with seizure preponderance

Correlation analyses between the ipsilateral hemispheric metrics and the proportion of wake-related seizures were run in theta and sigma bands (N_TLE_=21). The ipsilateral theta HE measured during wakefulness resulted to be positively correlated with the percentage of wake-related seizures (rho=0.51, p_uncorr_ 0.016, p_FDRcorr_= 0.063; Fig. 3a), while no correlation was found during sleep (Fig. 3b) nor in the sigma band.

**Fig 3:**
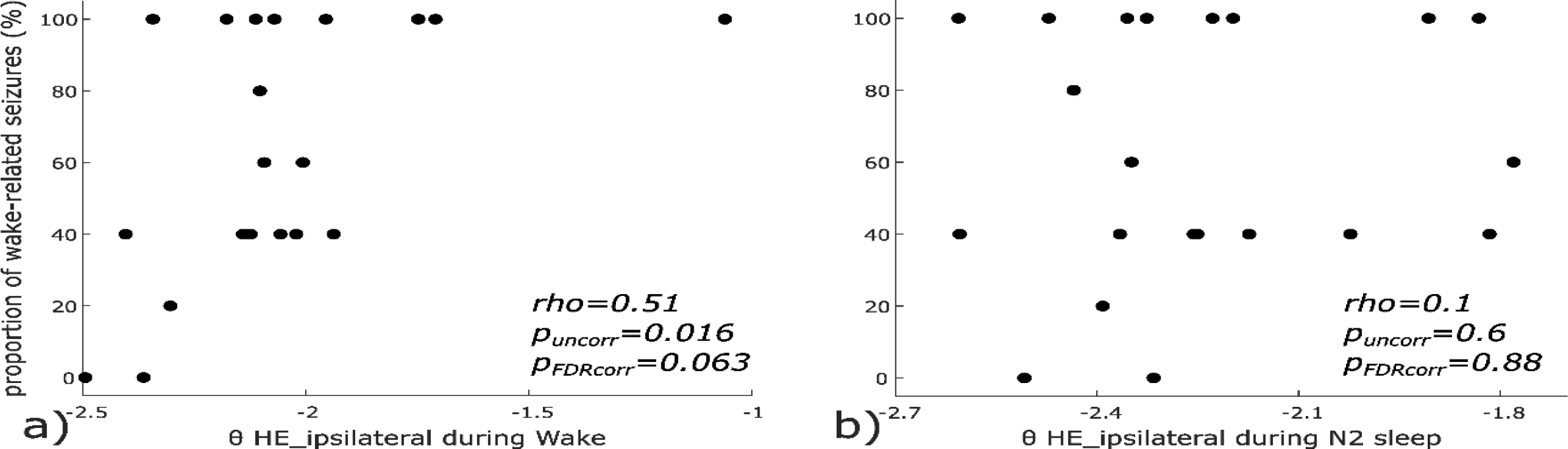
Scatterplot and results of Spearman correlation between the percentage of wake-related seizures and the theta hemispheric efficiency (HE) of the ipsilateral hemisphere during wakefulness (a) and N2 sleep (b). The percentage of wake-related seizures is displayed along the y-axis, where, for example, 60% indicate the 60-80% range.

### 3.3 Lobar network metrics

At the temporal-lobe level, we investigated whether the wake-to-sleep changes in network efficiency and clustering coefficient were lateralized toward the ipsilateral or contralateral temporal lobe (N_TLE_=21). We found significant changes of temporal-lobe segregation between wakefulness and sleep only in the ipsilateral hemisphere: the TCC decreased from wakefulness to N2 sleep in the delta band (z=2.6, p=.0434), but no correlation with the percentage of wake-related seizures was found.

### 3.4 Edge-wise analysis

As the multi-level analyses showed significant interactions in the theta band only, we ran NBS analyses in this frequency band (N_TLE_=21, N_HC_=13). The comparison of theta connectivity matrices during wakefulness between TLE and HC was the only one to hold a significant result (p=.004). Specifically, NBS identified a subnetwork comprising the ipsilateral hippocampus and parahippocampal gyrus that had stronger connections in TLE than in HC, during wakefulness (Fig. 4).

**Fig 4.**
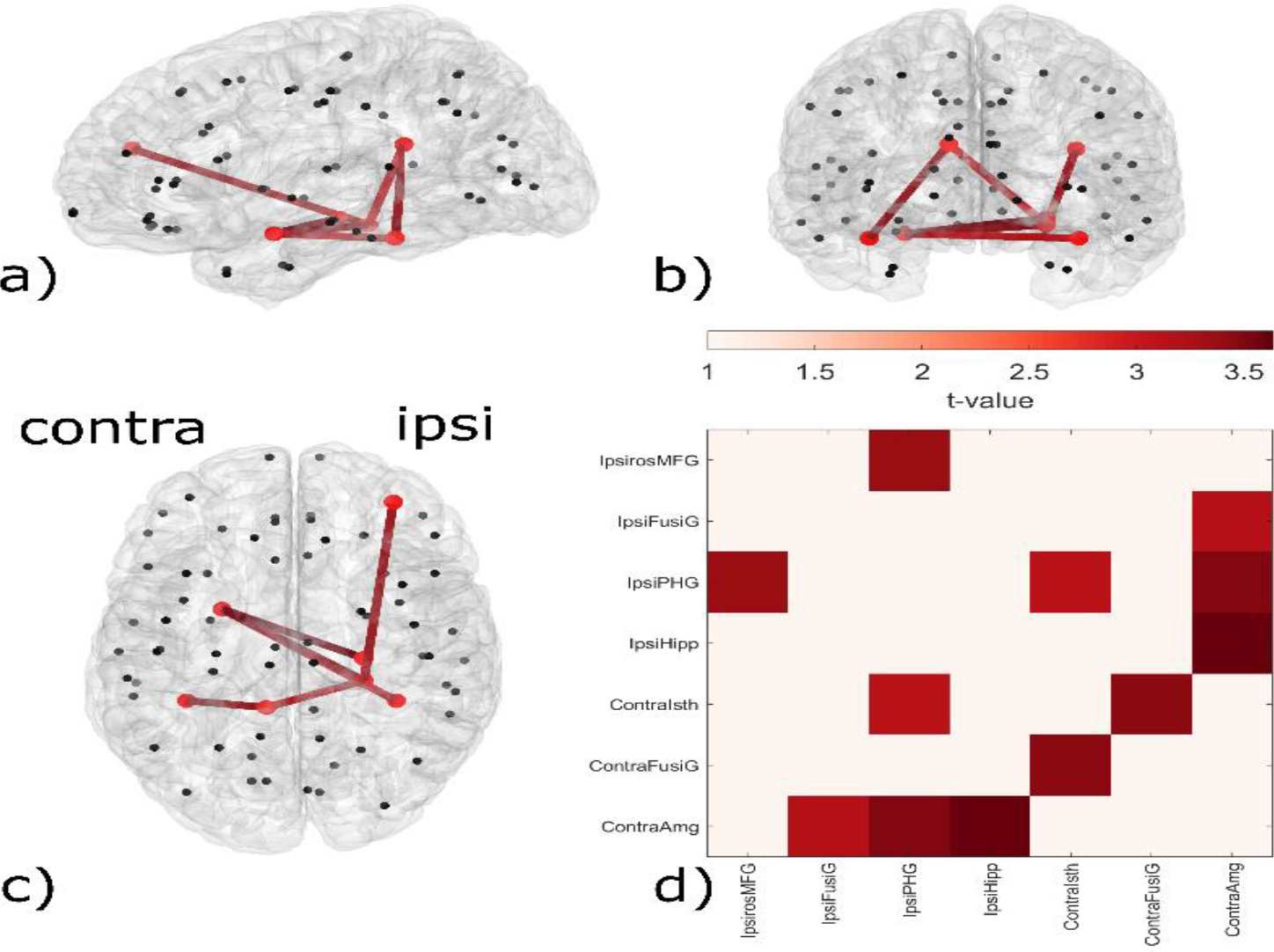
Representation of the network differences occurring in theta frequency band. a), b) and c): Cortical regions are represented as nodes of the network (black dots), while edges represent the connections between the regions (red lines). The figure highlights the differences between the brain network of TLE and HC during wakefulness: red lines constitute the subnetwork that in TLE is stronger than in HC. For these analyses, the networks were reorganized according to the ipsilateral and contralateral hemispheres: in the figure, the right hemisphere is the one ipsilateral to the epileptic focus. d): the figure depicts the names of the ROIs that belong to this subnetwork, where the intensity of the color reflects the supra-threshold t-value of each connection. Ipsi, ipsilateral; Contra, contralateral; MFG, middle frontal gyrus; FusiG, fusiform gyrus; PHG, parahippocampal gyrus; Hipp, hippocampus; Isth, isthmus; Amg, amygdala.

## 4 Discussion

### 4.1 Network alterations in TLE are detected in wakefulness but not in sleep

The significant interaction observed in theta frequency band shows higher GE in TLE patients than in HC during resting-state wakefulness, while such difference is not found during N2 sleep. Moreover, edge-wise analyses showed that a theta subnetwork with higher connectivity in patients (than in HC) is detectable only during wakefulness and comprises the ipsilateral hippocampus and parahippocampal gyrus, while no difference can be found between groups during N2. Additionally, a simple main effect of the group was found for 1) GE and avgCC in delta and 2) for avgCC in theta, showing that TLE patients’ brain networks at rest are not only more integrated but also more segregated than HCs’, irrespective of the vigilance state.

These results confirm and expand previous findings of pathological higher integration in TLE vs controls during wakefulness, which are further corroborated by the fact that a different connectivity metric was employed (Carboni et al., 2020; Duma et al., 2022). Since theta activity is known to often participate in inhibitory processes during wakefulness (Nir et al., 2017), the change in network configuration may indicate pathological inhibition, which will be discussed in more detail below. The other significant interaction observed in theta band shows that GE measured in patients is higher during wakefulness than sleep, which is difficult to interpret as other studies looking at generalized network changes in TLE during sleep are lacking.

Altogether, our results suggest that whole-brain network alterations between HC and TLE are detectable during wakefulness, while there is no observable difference during N2 sleep. Indeed, the above-mentioned pathological inhibition, visible during wakefulness (Nir et al., 2017), seems to be undetectable during sleep, where theta activity is more prominent.

### 4.2 Network metrics are altered in epilepsy during wake but not sleep

The interpretation that “pathological theta networks are visible during wakefulness but hidden by healthy theta activity during N2 sleep” is supported by the fact that the difference between wake and sleep is significant only for the ipsilateral hemispheric measures (theta HE and HCC) but not for the contralateral ones (Fig. 2). At the lobar level, however, this was not the case, implying that the affected network may extend beyond the temporal lobe, as also indicated by the theta inter-hemispheric subnetwork with stronger connections in TLE than HC (Fig. 4). Moreover, although not passing FDR correction, ipsilateral theta HE positively correlates with the proportion of wake-related seizures only during wakefulness and not during sleep (Fig 3). In other words, while the theta network appears to be pathological in awake TLE patients, it does not seem related to the disease during N2 sleep. The same holds true for delta band, where no significant interaction was found via ANOVA. Although not surviving multiple-comparison correction, the results suggest a correlation of global network metrics with the disease only during wakefulness (GE, avgCC), and not sleep. This suggests that the delta network, which has also been associated with cortical inhibition (Pigorini et al., 2015), is more strongly associated with the pathology during wakefulness than during sleep.

These results suggest that the wake state could more reliably inform us about the disease compared to sleep. This claim might seem controversial, as it is known that epileptic activity is more frequent during sleep with larger IED fields (Lambert et al., 2018; Malow et al., 1999, 1998). However, here we purposely selected EEG epochs not containing interictal epileptic activity, to study a “negative EEG” scenario. Given that IEDs are more easily detected during N2, more cortical spikes could have gone undetected in the wake epochs compared to the sleep epochs. These ‘hidden’ spikes could have contributed to the difference between groups during wakefulness, while sleep epochs may have been better ‘cleared’ of IEDs. Altogether, our results indicate that, in absence of IEDs and seizures, it could be easier to distinguish a physiologic brain from a pathologic one by looking at EEG-derived brain networks during wakefulness rather than N2 sleep.

### 4.3 Combined higher integration and segregation exist in TLE

Regarding the nature of these network changes, it is interesting to notice that GE and avgCC (and also HE and HCC) have similar patterns. In other words, brain integration and brain segregation do not seem to be mutually exclusive, but rather two faces of a coin, that co-exist and change simultaneously. Networks that are highly clustered (high avgCC) yet have characteristically short path lengths (high GE) are usually referred to as “small-world” (Watts and Strogatz, 1998). This configuration is characteristic of healthy functioning networks and is thought to facilitate neural synchronization (Ferri et al., 2007; Stam, 2004; Watts and Strogatz, 1998). In epilepsy, a brain network with a “small world” configuration was related to seizure generation (Netoff et al., 2004; Percha et al., 2005). During ictal discharges, the network exhibits this type of organization, while during interictal discharges, the network appears to be more randomly organized (Ponten et al., 2007). A study found an increase of both avgCC and GE even during interictal activity -without exclusion of IEDs-in patients with generalized epilepsy (Chavez et al., 2010). We observed similar trends for both network metrics at low frequencies, suggesting that this may also apply to pathologic networks during resting state EEG, free from scalp-visible interictal epileptiform activity. This further suggests that epileptic brain networks are not exclusively unbalanced toward more integrated systems (Carboni et al., 2020), but are rather characterized by both increased segregation and integration.

### 4.4 Network changes in low frequencies suggest increased inhibition in TLE

Another interesting aspect is that epilepsy-related network changes were found in the lower part of the frequency spectrum (delta and theta bands) and not at higher frequencies. Indeed, the main effect in sigma band reflects a difference related to the vigilance state -likely to be ascribed to sleep spindles (De Gennaro and Ferrara, 2003)-indicating higher integration and segregation during N2 sleep across both groups. The absence of correlation between the seizure preponderance and any network metric in sigma band (global and hemispheric) is yet another factor speaking in favor of this interpretation. Importantly there was no one-to-one correspondence between the power of the signal and the network metrics across frequency bands (see Appendix), suggesting that network analyses remain unbiased by the signal power and provide independent information.

Physiologically, delta is fundamental for large-scale cortical integration (Bruns and Eckhorn, 2004) and theta is a dominant hippocampal rhythm (Kahana et al., 1999). As already mentioned, theta activity was documented in the waking state of healthy individuals and was thought to reflect local functional inhibition (neuronal ‘off’ periods) (Nir et al., 2017). Delta and theta waves are also a normal characteristic of N2 sleep stage, occupying almost 20% of the epochs (Chokroverty, 2009). Moreover, slow wave activity in awake adults is a well-recognized indicator of pathologic activity and cerebral dysfunction (Britton et al., 2016). In these terms, a greater theta GE and higher connectivity in patients than in controls could reflect pathological functional inhibition, especially in the ipsilateral hippocampus and parahippocampal gyrus (NBS results show higher connectivity in TLE, see Fig 4). Therefore, increased connectivity of inhibitory networks could be a marker of compensatory mechanisms in the context of epileptic activity rather than the marker of epileptic activity per se.

Although we cannot distinguish between excitatory and inhibitory connections based on our connectivity analysis, recent intracranial EEG studies suggest that the main mechanism active during resting state in TLE is inhibitory. According to the Interictal Suppression Hypothesis, seizures are prevented during interictal activity by successful inhibition of the epileptogenic zone (EZ) (Johnson et al., 2023). To this regard, it was shown that regions belonging to the EZ were indeed those with the highest inward connections (sinks) (Gunnarsdottir et al., 2021; Johnson et al., 2023), likely reflecting inhibitory inputs from regions external to the EZ, that could be replaced by outward connections supporting seizure spread once the seizure begins (Narasimhan et al., 2020). High functional segregation of the EZ was also shown at rest (Bandt et al., 2014; Johnson et al., 2023; Schevon et al., 2012). Even if we cannot distinguish between inward and outward connections, we can speculate that the widespread integration seen at the low frequencies mainly has an inhibitory role which, together with high level of segregation isolating the EZ, could represent a tentative control of the epileptic activity. Our previous studies suggested that the increased integration observed in patients was indicative of a wider pathological network within the brain that supported a greater spread of epileptic activity (Carboni et al., 2020, 2019). With this study, we reiterate the concept and speculate that, considering the concurrent segregation increase, this more integrated and segregated network may have a compensatory interictal inhibitory role.

### 4.5 Limitations

The main limitations comprise the use of a template head model for source reconstruction instead of individual head models (because MRI was not available for HC), missing wake EEG for 10 HC, and, overall, the fact that EEG of TLE patients and HC was recorded at different sites and under different circumstances, *i.e*. in an epilepsy monitoring unit and a sleep laboratory. Sleep EEG of HC was consistently recorded at night whereas daytime naps were also considered for TLE patients. This renders our results specific for N2 but not for nighttime sleep or even a specific part of the night, so that effects of the circadian clock on the epileptic network (Karoly et al., 2021) were not regarded. Wake EEG of HC was taken from eyes-open segments while EEG of TLE patients was taken most often from closed-eyes segments, which might have introduced more variability in the alpha-band spectral power. However, given the absence of connectivity changes found in alpha band, this confound does not affect the interpretation of our results. Lastly, TLE patients were treated with antiseizure medications which may confound spectral power although spectral power and graph measures seem mostly unrelated in our study (see Fig S2).

## 5 Conclusions

Altogether, our results suggest global integration and global segregation increase in TLE, suggesting a pathological tendency toward a ‘small world’ configuration. These global network alterations were observed, both during wakefulness and N2 sleep, mainly in the lower part of the frequency spectrum (delta and theta). In the interictal state, high functional integration and segregation might play a protective inhibitory role against epileptic activity.

During N2 sleep, physiologic theta waves and potentially ‘purer’ spike-free epochs may complicate the detection of pathological connectivity patterns suggesting that, in absence of IEDs, it might be easier to detect epilepsy-related network alterations during wakefulness than during sleep.

## Data Availability

All patients agreed to data reuse for research purposes, but not all agreed to data sharing. Therefore, the EEG data used in this study cannot be shared on a public platform.

## Authors contributions

**IR**: Conceptualization; Data curation; Methodology; Formal analysis; Interpretation; Writing -original draft; Writing - review & editing. **BV**: Conceptualization; Patient selection; Data curation; Interpretation; Writing - original draft; Writing - review & editing. **MC**: Data curation; Methodology; Formal analysis; Writing - review & editing. **NR**: Methodology; Writing - review & editing. **LS**: Resources; Writing - review & editing. **GT**: Funding acquisition; Writing - review & editing. **MS**: Resources; Writing - review & editing. **LP**: Conceptualization; Data curation; Interpretation; Writing - review & editing. **SV**: Conceptualization; Interpretation; Supervision; Funding acquisition; Writing - original draft; Writing - review & editing.

## References

Arbune AA, Popa I, Mindruta I, Beniczky S, Donos C, Daneasa A, et al. Sleep modulates effective connectivity: A study using intracranial stimulation and recording. Clin Neurophysiol 2020;131:529–41. 10.1016/j.clinph.2019.09.010.

Bandt SK, Bundy DT, Hawasli AH, Ayoub KW, Sharma M, Hacker CD, et al. The role of resting state networks in focal neocortical seizures. PLoS One 2014;9:1–10. 10.1371/journal.pone.0107401.

Bazil CW. Seizure modulation by sleep and sleep state. Brain Res 2019;1703:13–7. 10.1016/j.brainres.2018.05.003.

Berg AT, Berkovic SF, Brodie MJ, Buchhalter J, Cross JH, Van Emde Boas W, et al. Revised terminology and concepts for organization of seizures and epilepsies: Report of the ILAE Commission on Classification and Terminology, 2005-2009. Epilepsia 2010;51:676–85. 10.1111/j.1528-1167.2010.02522.x.

Bosak M, Słowik A, Kacorzyk R, Turaj W. Implementation of the new ILAE classification of epilepsies into clinical practice — A cohort study. Epilepsy Behav 2019;96:28–32. 10.1016/j.yebeh.2019.03.045.

Britton J, Frey L, Hopp J, Korb P, Koubeissi M, Lievens W, et al. Electroencephalography (EEG): An Introductory Text and Atlas of Normal and Abnormal Findings in Adults, Children, and Infants [Internet]. Chicago: American Epilepsy Society; 2016.; 2016.

Brunet D, Murray MM, Michel CM. Spatiotemporal analysis of multichannel EEG: CARTOOL. Comput Intell Neurosci 2011;2011. 10.1155/2011/813870.

Bruns A, Eckhorn R. Task-related coupling from high-to low-frequency signals among visual cortical areas in human subdural recordings. Int J Psychophysiol 2004;51:97–116. 10.1016/j.ijpsycho.2003.07.001.

Cammoun L, Gigandet X, Meskaldji D, Thiran JP, Sporns O, Do KQ, et al. Mapping the human connectome at multiple scales with diffusion spectrum MRI. J Neurosci Methods 2012;203:386–97. 10.1016/j.jneumeth.2011.09.031.

Carboni M, Rubega M, Iannotti GR, De Stefano P, Toscano G, Tourbier S, et al. The network integration of epileptic activity in relation to surgical outcome. Clin Neurophysiol 2019;130:2193–202. 10.1016/j.clinph.2019.09.006.

Carboni M, De Stefano P, Vorderwülbecke BJ, Tourbier S, Mullier E, Rubega M, et al. Abnormal directed connectivity of resting state networks in focal epilepsy. NeuroImage Clin 2020;27:102336. 10.1016/j.nicl.2020.102336.

Chavez M, Valencia M, Navarro V, Latora V, Martinerie J. Functional modularity of background activities in normal and epileptic brain networks. Phys Rev Lett 2010;104:1–4. 10.1103/PhysRevLett.104.118701.

Chokroverty S. An overview of normal sleep. Sleep Disord. Med. Basic Sci. Tech. considerations, Clin. Asp. 3rd ed., Philadelphia: Saunders Elsevier; 2009, p. 5–21.

Coito A, Genetti M, Pittau F, Iannotti GR, Thomschewski A, Höller Y, et al. Altered directed functional connectivity in temporal lobe epilepsy in the absence of interictal spikes: A high density EEG study. Epilepsia 2016;57:402–11. 10.1111/epi.13308.

Duma GM, Danieli A, Mattar MG, Baggio M, Vettorel A, Bonanni P, et al. Resting state network dynamic reconfiguration and neuropsychological functioning in temporal lobe epilepsy: An HD-EEG investigation. Cortex 2022;157:1–13. 10.1016/j.cortex.2022.08.010.

Ferri R, Rundo F, Bruni O, Terzano MG, Stam CJ. Small-world network organization of functional connectivity of EEG slow-wave activity during sleep. Clin Neurophysiol 2007;118:449–56. 10.1016/j.clinph.2006.10.021.

Fisher RS, Acevedo C, Arzimanoglou A, Bogacz A, Cross JH, Elger CE, et al. ILAE Official Report: A practical clinical definition of epilepsy. Epilepsia 2014;55:475–82. 10.1111/epi.12550.

Fisher RS, Cross JH, French JA, Higurashi N, Hirsch E, Jansen FE, et al. Operational classification of seizure types by the International League Against Epilepsy: Position Paper of the ILAE Commission for Classification and Terminology. Epilepsia 2017;58:522–30. 10.1111/epi.13670.

De Gennaro L, Ferrara M. Sleep spindles: An overview. Sleep Med Rev 2003;7:423–40. 10.1053/smrv.2002.0252.

Gunnarsdottir KM, Li A, Smith RJ, Kang J-Y, Korzeniewska A, Crone NE, et al. Source-sink connectivity: A novel interictal EEG marker for seizure localization. Brain 2021;145:2021.10.15.464594. 10.1093/brain/awac300.

Hagmann P, Cammoun L, Gigandet X, Meuli R, Honey CJ, Van Wedeen J, et al. Mapping the structural core of human cerebral cortex. PLoS Biol 2008;6:1479–93. 10.1371/journal.pbio.0060159.

Holmes C, Hoge R, Collins L, Woods R, Toga AW, Evans AC. Enhancement of MR Images Using Registration for Signal Averaging. J Comput Assist Tomogr 1998;22:324–33.

Iber C, Ancoli-Israel S, Chesson A, Quan S. The AASM Manual for the Scoring of Sleep and Associated Events 2007.

Jain S V., Kothare S V. Sleep and Epilepsy. Semin Pediatr Neurol 2015;22:86–92. 10.1016/j.spen.2015.03.005.

Johnson GW, Doss DJ, Morgan VL, Paulo DL, Cai LY, Shless JS, et al. The Interictal Suppression Hypothesis in focal epilepsy: network-level supporting evidence. Brain 2023;146:2828–45. 10.1093/brain/awad016.

Kahana MJ, Sekuler R, Caplan JB, Kirschen M, Madsen JR. Human theta oscillations exhibit task dependence during virtual maze navigation. Nature 1999;399:781–4. 10.1038/21645.

Karoly PJ, Rao VR, Gregg NM, Worrell GA, Bernard C, Cook MJ, et al. Cycles in epilepsy. Nat Rev Neurol 2021;17:267–84. 10.1038/s41582-021-00464-1.

Lagarde S, Roehri N, Lambert I, Trebuchon A, McGonigal A, Carron R, et al. Interictal stereotactic-EEG functional connectivity in refractory focal epilepsies. Brain 2018;141:2966–80. 10.1093/brain/awy214.

Lambert I, Roehri N, Giusiano B, Carron R, Wendling F, Benar C, et al. Brain regions and epileptogenicity influence epileptic interictal spike production and propagation during NREM sleep in comparison with wakefulness. Epilepsia 2018;59:235–43. 10.1111/epi.13958.

Latora V, Marchiori M. Efficient behavior of small-world networks. Phys Rev Lett 2001;87:198701-1-198701–4. 10.1103/PhysRevLett.87.198701.

Malow BA, Lin X, Kushwaha R, Aldrich MS. Interictal spiking increases with sleep depth in temporal lobe epilepsy. Epilepsia 1998;39:1309–16. 10.1111/j.1528-1157.1998.tb01329.x.

Malow BA, Selwa LM, Ross D, Aldrich MS. Lateralizing value of interictal spikes on overnight sleep-EEG studies in temporal lobe epilepsy. Epilepsia 1999;40:1587–92. 10.1111/j.1528-1157.1999.tb02044.x.

Michel CM, Brunet D. EEG source imaging: A practical review of the analysis steps. Front Neurol 2019;10. 10.3389/fneur.2019.00325.

van Mierlo P, Papadopoulou M, Carrette E, Boon P, Vandenberghe S, Vonck K, et al. Functional brain connectivity from EEG in epilepsy: Seizure prediction and epileptogenic focus localization. Prog Neurobiol 2014;121:19–35. 10.1016/j.pneurobio.2014.06.004.

Narasimhan S, Kundassery KB, Gupta K. Seizure-onset regions demonstrate high inward directed connectivity during resting-state: An SEEG study in focal epilepsy. Epilepsia 2020;61:2534–2544. 10.1111/epi.16686.Seizure-onset.

Netoff TI, Clewley R, Arno S, Keck T, White JA. Epilepsy in small-world networks. J Neurosci 2004;24:8075–83. 10.1523/JNEUROSCI.1509-04.2004.

Nir Y, Andrillon T, Marmelshtein A, Suthana N, Cirelli C, Tononi G, et al. Selective neuronal lapses precede human cognitive lapses following sleep deprivation. Nat Med 2017;23:1474–80. 10.1038/nm.4433.

Oostenveld R, Fries P, Maris E, Schoffelen JM. FieldTrip: Open source software for advanced analysis of MEG, EEG, and invasive electrophysiological data. Comput Intell Neurosci 2011;2011. 10.1155/2011/156869.

Percha B, Dzakpasu R, Zochowski M, Parent J. Transition from local to global phase synchrony in small world neural network and its possible implications for epilepsy. Phys Rev E - Stat Nonlinear, Soft Matter Phys 2005;72:1–6. 10.1103/PhysRevE.72.031909.

Perogamvros L, Baird B, Seilbold M, Riedner B, Boly M, Tononi G. The Phenomenal Contents and Neural Correlates of Spontaneous Thoughts across Wakefulness, NREM Sleep, and REM Sleep. J Cogn Neurosci 2017;29:10:1766–77. 10.1162/jocn.

Pigorini A, Sarasso S, Proserpio P, Szymanski C, Arnulfo G, Casarotto S, et al. Bistability breaks-off deterministic responses to intracortical stimulation during non-REM sleep. Neuroimage 2015;112:105–13. 10.1016/j.neuroimage.2015.02.056.

Ponten SC, Bartolomei F, Stam CJ. Small-world networks and epilepsy: Graph theoretical analysis of intracerebrally recorded mesial temporal lobe seizures. Clin Neurophysiol 2007;118:918–27. 10.1016/j.clinph.2006.12.002.

Reuter M, Schmansky NJ, Rosas HD, Fischl B. Within-subject template estimation for unbiased longitudinal image analysis. Neuroimage 2012;61:1402–18. 10.1016/j.neuroimage.2012.02.084.

Rubega M, Carboni M, Seeber M, Pascucci D, Tourbier S, Toscano G, et al. Estimating EEG Source Dipole Orientation Based on Singular-value Decomposition for Connectivity Analysis. Brain Topogr 2019;32:704–19. 10.1007/s10548-018-0691-2.

Rubinov M, Sporns O. Complex network measures of brain connectivity: Uses and interpretations. Neuroimage 2010;52:1059–69. 10.1016/j.neuroimage.2009.10.003.

Sammaritano M, Gigli GL, Gotman J. Interictal spiking during wakefulness and sleep and the localization of foci in temporal lobe epilepsy. Neurology 1991;41.

Schevon CA, Weiss SA, McKhann G, Goodman RR, Yuste R, Emerson RG, et al. Evidence of an inhibitory restraint of seizure activity in humans. Nat Commun 2012;3:1011–60. 10.1038/ncomms2056.

Siclari F, Baird B, Perogamvros L, Bernardi G, LaRocque JJ, Riedner B, et al. The neural correlates of dreaming. Nat Neurosci 2017;20:872–8. 10.1038/nn.4545.

Slinger G, Otte WM, Braun KPJ, van Diessen E. An updated systematic review and meta-analysis of brain network organization in focal epilepsy: Looking back and forth. Neurosci Biobehav Rev 2021. 10.1016/j.neubiorev.2021.11.028.

Staljanssens W, Strobbe G, Van Holen R, Keereman V, Gadeyne S, Carrette E, et al. EEG source connectivity to localize the seizure onset zone in patients with drug resistant epilepsy. NeuroImage Clin 2017;16:689–98. 10.1016/j.nicl.2017.09.011.

Stam CJ. Functional connectivity patterns of human magnetoencephalographic recordings: A “small-world” network? Neurosci Lett 2004;355:25–8. 10.1016/j.neulet.2003.10.063.

Tourbier S, Aleman-Gomez Y, Griffa A, Bach Cuadra M, Hagmann P. connectomicslab/connectomemapper3: Connectome Mapper v3.0.0-beta-20200206 2020. 10.5281/ZENODO.3653749.

Usami K, Matsumoto R, Kobayashi K, Hitomi T, Shimotake A, Kikuchi T, et al. Sleep modulates cortical connectivity and excitability in humans: Direct evidence from neural activity induced by single-pulse electrical stimulation. Hum Brain Mapp 2015;36:4714–29. 10.1002/hbm.22948.

Varatharajah Y, Joseph B, Brinkmann B, Morita-Sherman M, Fitzgerald Z, Vegh D, et al. Quantitative analysis of visually reviewed normal scalp EEG predicts seizure freedom following anterior temporal lobectomy. Epilepsia 2022:1630–42. 10.1111/epi.17257.

Verhoeven T, Coito A, Plomp G, Thomschewski A, Pittau F, Trinka E, et al. Automated diagnosis of temporal lobe epilepsy in the absence of interictal spikes. NeuroImage Clin 2018;17:10–5. 10.1016/j.nicl.2017.09.021.

Vorderwülbecke BJ, Carboni M, Tourbier S, Brunet D, Seeber M, Spinelli L, et al. High-density Electric Source Imaging of interictal epileptic discharges: How many electrodes and which time point? Clin Neurophysiol 2020;131:2795–803. 10.1016/j.clinph.2020.09.018.

Watts DJ, Strogatz SH. Collective dynamics of ‘small-world’ networks. Nature 1998;393:440–2.

Yuan X, Sun M. The value of rapid eye movement sleep in the localization of epileptogenic foci for patients with focal epilepsy. Seizure 2020;81:192–7. 10.1016/j.seizure.2020.06.009.

