## Appendix for "Network alterations in temporal lobe epilepsy during non-rapid eye movement sleep and wakefulness"

### Appendix
#### Supplementary Methods

##### Network metrics calculation

The GE reflects how well information is propagated over distributed brain regions and is a measure of networks' *integration*: GE is related to the "shortest path length" or, more simply, the "distance". In the case of weighted undirected networks like here, the distance $\mathrm{d}_{ij}^{w}$ between node $i$ and node $j$ is calculated as the sum of the inverse of the strongest weights that directly connect $i$ to $j$. In other words the shortest path length is the length of the path with the least resistance to information transfer. Intuitively, the higher the distance between $i$ and $j$, the smaller the efficiency in their communication. In this sense, the efficiency $E_{ij}$ of the communication between $i$ and $j$ can be defined as inversely proportional to their distance: $E_{ij} = 1/\mathrm{d}_{ij}^{w}$ (Latora and Marchiori, 2001). The average efficiency of the network, GE, can then be calculated as the average inverse shortest path length of the network (Latora and Marchiori, 2001; Rubinov and Sporns, 2010):

$$GE = \frac{1}{n}\sum_{i\in N}\frac{\sum_{j\in N, j\neq i} E_{ij}}{n-1} = \frac{1}{n(n-1)}\sum_{i\neq j\in N}\frac{1}{\mathrm{d}_{ij}^{w}}$$

The hemispheric efficiency (HE) was computed as the GE of the network containing only the nodes of one hemisphere and their intra-hemispheric connections. Similarly, the temporal lobe efficiency (TE) was computed as the GE of the temporal lobe network.

The second metric, the avgCC, reflects the preponderance of clustered connectivity around individual nodes and therefore reflects the networks' *segregation*. It is computed as the average CC of a network's nodes. The CC of a node is obtained as the fraction of the node's neighbors that are also neighbors of each other –as measured by the "number of triangles around the node itself"- (Watts and Strogatz, 1998). Since it reflects the tendency of the nearest neighbors of the node to be interconnected with each other, it reflects the tendency of the network of clustering around individual nodes. More formally, avgCC is calculated as:

$$avgCC = \frac{1}{n}\sum_{i\in N} CC_i = \frac{1}{n}\sum_{i\in N}\frac{2t_i^{w}}{k_i(k_i-1)}$$

where $k_i$ is the degree of the node and $t_i^{w}$ is the number of triangles attached to the node that, for weighted networks, is calculated as the weighted geometric mean of triangles around the node $i$ (Onnela et al., 2005; Rubinov and Sporns, 2010):

$$t_i^{w} = \frac{1}{2}\sum_{j,h\in N}(w_{ij}w_{ih}w_{hj})^{1/3}$$

The hemispheric clustering coefficient (HCC) was computed as the average CC of the network containing only the nodes belonging to one hemisphere and their intra-hemispheric connections. Similarly, the temporal lobe CC (TCC) was computed as the average CC of the temporal lobe network.

##### Edge-wise analyses

The network-based statistic (NBS) method was used to identify any subnetwork that significantly differed between the two groups compared. To reduce the number of tests, this was done only for the frequency bands where a significant interaction was found for the global network metrics (GE or avgCC). For each of these frequency bands, we ran four comparisons (TLE_W vs. HC_W; TLE_N2 vs. HC_N2; TLE_W vs. TLE_N2 and HC_W vs. HC_N2), where we tested the hypothesis that "group 1 has stronger –or weaker– connections than group 2". T-tests were used for mass univariate testing and a t-value of 3.1 was used as threshold to identify connected graph components. Then, 5000 permutations were used to define the significance of such components. The network-based statistic (NBS) method was chosen to control the family-wise error rate (Zalesky et al., 2010). In practice, we retained patients with temporal lobe epilepsy but not bilateral (N=21, 8/21=38% LTLE) and reorganized the connectivity matrices into ipsilateral and contralateral to the epileptogenic side (rather than left or right hemisphere). We flipped right and left hemispheres in the same proportion of healthy controls, depending on the comparison:1) in TLE_W vs. HC_W (NTLE,W=21 and NHC,W=13), the hemispheres were flipped in 5/13 (38%) of HC; 2) in TLE_N2 vs. HC_N2 (NTLE,N2=21 and NHC,N2=21), the hemispheres were flipped in 8/21 (38%) HC. Hemispheres were not flipped when comparing HC_W vs. HC_N2 (NHC,N2=13 and NHC,W=13). The obtained p-values were further Bonferroni-corrected to account for the 4 tests ran for each frequency band.

##### Power spectral density

The power spectral density (PSD) was calculated between 1-40 Hz using multiple tapers. The chosen spectral smoothing box of 2 Hz yielded three tapers. The relative power in each frequency band was calculated for every ROI as the percentage of the power in the specific band with respect to the total power, which were both calculated with the 'trapz' Matlab function. For each band, the relative power was averaged over ROI and used for supplementary correlation analyses. We calculated the correlation between each graph measure (GE and avgCC) and the relative power in each frequency band (5), for each group (4). We used Bonferroni correction to account for the 20 tests performed for each graph measure. Pearson's or Spearman's correlations were run depending on the presence of outliers, which were automatically detected.

#### Supplementary Results

##### Hemispheric changes in healthy controls

***Hemispheric efficiency*** Wilcoxon tests, run on the HE of healthy controls, showed no significant change of the network metric from wake to N2 sleep neither in the "ipsilateral" hemisphere, nor in the "contralateral" hemisphere (see Fig. S1a).

***Hemispheric clustering coefficient*** Similarly, Wilcoxon tests, run on the HCC of healthy controls, showed no significant change of the network metric from wake to N2 sleep neither in the "ipsilateral" hemisphere, nor in the "contralateral" hemisphere (see Fig. S1b).

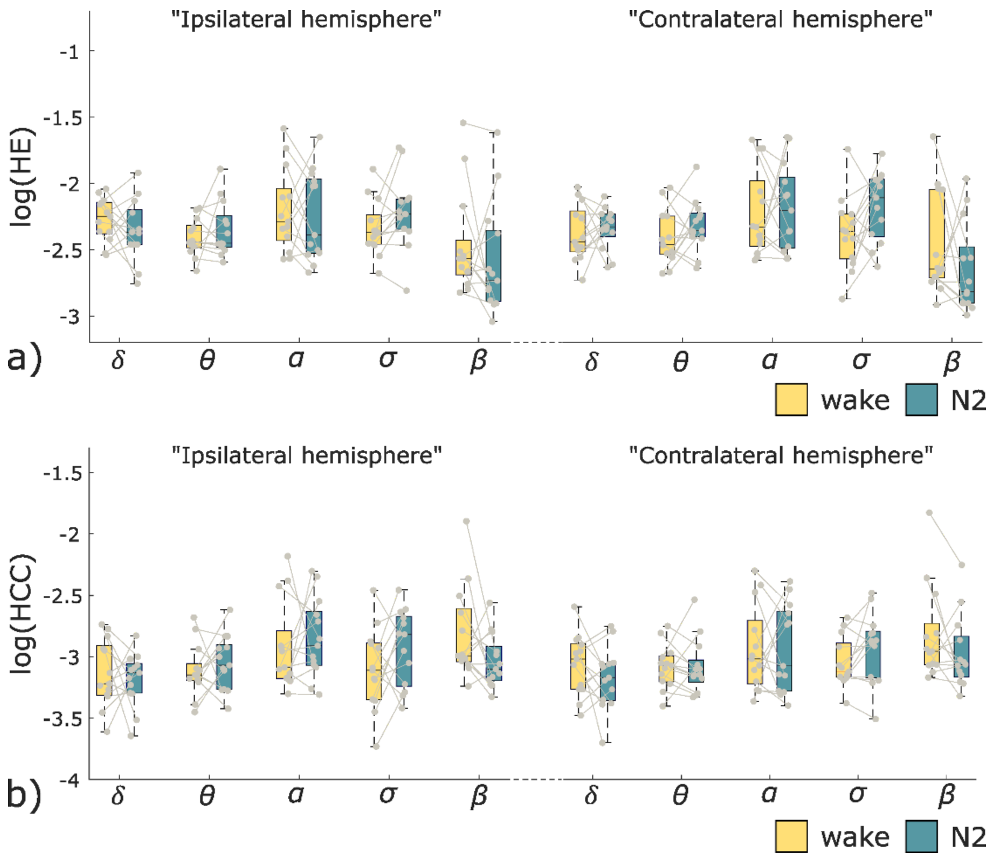


**Fig S1:** Boxplots of the a) hemispheric efficiency and b) hemispheric clustering coefficient measured in HC subjects across frequency bands during wakefulness and N2 sleep. The right and left hemispheres were flipped in half of the controls.

**Temporal lobe changes in HC**

Wilcoxon tests, run on the temporal lobe efficiency (TE) of healthy controls, showed no significant change of the network metric from wake to N2 sleep neither in the “ipsilateral” temporal lobe nor in the “contralateral” temporal lobe. Similarly, the temporal CC (TCC) did not show significant changes in any frequency band from wake to N2 sleep, neither in the “ipsilateral” temporal lobe nor in the “contralateral” temporal lobe.

**Correlation of signal’s power with network metrics**

A significant positive correlation was found between both graph measures and the relative power in theta band (GE: rho=0.7, p=.0058; avgCC: rho=0.62, p=.04, see Fig. S2) and in alpha band (GE: rho=0.77, p=0.040; avgCC: rho=0.83, p=.01) only in HC. In TLE patients, no correlation was found between network metrics and signal’s power.

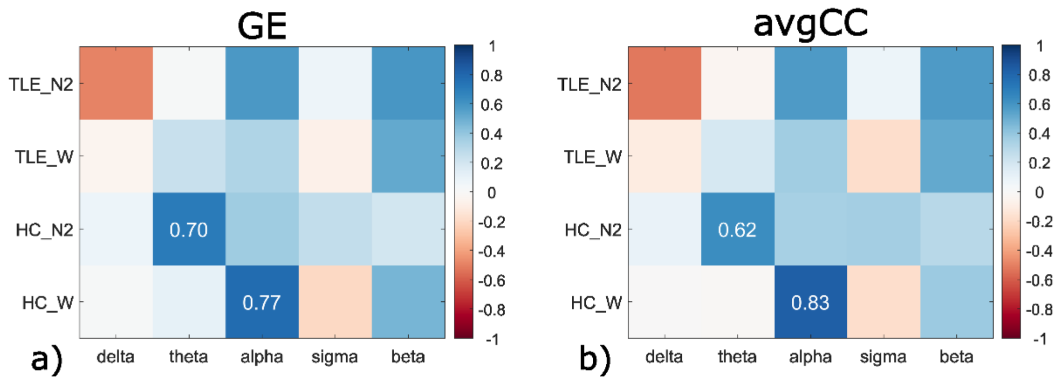


**Fig S2**: Results of correlation analysis between relative power and: a) GE; b) avgCC. Analyses are run for each group (TLE and HC, in wakefulness W and N2 sleep). The colormap represents correlation values between the specific graph measure and the relative power in the specific frequency band, for each group. The correlation values are reported only for significant correlations ($p<.05$).

### References

Latora V, Marchiori M. Efficient behavior of small-world networks. Phys Rev Lett 2001;87:198701-1-198701–4. https://doi.org/10.1103/PhysRevLett.87.198701.

Onnela JP, Saramäki J, Kertész J, Kaski K. Intensity and coherence of motifs in weighted complex networks. Phys Rev E - Stat Nonlinear, Soft Matter Phys 2005;71:1–4. https://doi.org/10.1103/PhysRevE.71.065103.

Rubinov M, Sporns O. Complex network measures of brain connectivity: Uses and interpretations. Neuroimage 2010;52:1059–69. https://doi.org/10.1016/j.neuroimage.2009.10.003.

Watts DJ, Strogatz SH. Collective dynamics of 'small-world' networks. Nature 1998;393:440–2.

Zalesky A, Fornito A, Bullmore ET. Network-based statistic: Identifying differences in brain networks. Neuroimage 2010;53:1197–207. https://doi.org/10.1016/j.neuroimage.2010.06.041.
